# Development of novel clinical examination scales for the measurement of disease severity in Creutzfeldt-Jakob disease

**DOI:** 10.1101/2020.11.01.20224089

**Authors:** Akin Nihat, Tze How Mok, Hans Odd, Andrew Thompson, Diana Caine, Kirsty McNiven, Veronica O’Donnell, Selam Tesfamichael, Peter Rudge, John Collinge, Simon Mead

## Abstract

**Objective:** Sporadic Creutzfeldt-Jakob disease (sCJD) causes rapidly-progressive dementia and complex abnormalities of motor systems with striking phenotypic heterogeneity, but no tools are available for the clinician to determine disease severity from bedside cognitive and neurological assessments. We used a robust statistical methodology and routinely-collected examination data to develop and validate short clinical rating scales quantifying longitudinal motor and cognitive dysfunction in sCJD.

**Methods:** We undertook a retrospective analysis of clinical examination data from the prospective National Prion Monitoring Cohort study, October 2008 – December 2016. Rasch analysis was used to iteratively construct interval scales measuring composite cognitive and motor dysfunction from pooled bedside neurological and cognitive examination tests.

A longitudinal clinical examination dataset was constructed from a total of 528 patients with sCJD, comprising 1030 Motor Scale and 757 Cognitive Scale scores, over 130 patient-years of study, and used to demonstrate scale utility.

**Results:** The Rasch-derived Motor Scale consists of 8 items, including examination items reliant on pyramidal, extrapyramidal and cerebellar systems. The Cognitive Scale comprises 6 items, and includes measures of executive function, language, visual perception and memory. Both scales are unidimensional, perform consistently regardless of age or gender and have excellent inter-rater reliability. Each scale can be completed in a few minutes at the bedside, as part of a normal neurocognitive examination. Several uses of the scales, in measuring longitudinal change, prognosis, and phenotypic heterogeneity are illustrated.

**Interpretation:** These two novel scales measuring motor and cognitive dysfunction in sCJD should prove useful to objectively measure phenotypic and clinical change in future clinical trials and for patient stratification. This statistical approach can help to overcome obstacles to assessing clinical change in rapidly-progressive, multisystem conditions with limited longitudinal follow-up.

## Introduction

The human prion diseases are a group of universally fatal neurodegenerative conditions that share a common mechanism: the autocatalytic, templated misfolding of the constitutively expressed prion protein (PrP^C^) into disease-related multimeric assemblies including protease-resistant forms designated PrP^Sc 1^. They can be separated aetiologically into sporadic (approximately 85% of incident cases), inherited (10-15%) and acquired (<1%) subtypes. The median clinical duration from symptom onset in sporadic Creutzfeldt-Jakob disease (sCJD) is 4 months, although disease courses ranging from short weeks to several years are recognised ^2, 3^. Even within aetiological groups there is enormous heterogeneity in clinical phenotype ^3-5^, duration and histopathology, partly reflecting the propagation of distinct conformational strains of misfolded prion protein ^1^.

Well-designed clinical rating scales offer quantitative measures of disease-specific change applicable to clinical trials as eligibility or stratification criteria, outcome measures, or for prognostic modelling. They may also provide useful information for patients, caregivers and healthcare professionals about relevant aspects of disease progression, specific functions or symptoms. In 2013 we proposed the Medical Research Council Prion Disease Rating Scale (MRC Scale) ^6^, a validated functional outcome measure of disease progression in sporadic CJD, acquired by brief carer interview. The single MRC Scale score encapsulates all aspects of a patient’s performance, and allows direct group and individual comparison of disease progression in a clinically and statistically meaningful manner ^7^, and usefully, can be acquired remotely. Weaknesses of the MRC Scale are that it does not directly measure the impairments that contribute to functional deficits, and that it relies on a witness report.

Here, we sought to develop physician examination-orientated neurological and cognitive rating scales to complement the functionally-oriented MRC Scale as clinimetric tools. In doing so, we demonstrate that simple clinical data collected routinely as part of a bedside neurological and cognitive examination in a well-defined cohort, can be used to construct meaningful disease and system-specific clinical rating scales with robust statistical properties. These new tools in prion disease have multiple potential uses in clinical trials, research, and patient and family feedback. This offers a model to develop similar tools in other complex dementia syndromes.

## Methods

### Study population

Details of the National Prion Monitoring Cohort (NPMC) schedule have been published ^6^. At enrolment to the NPMC, each patient has a comprehensive clinical assessment. This includes a standardised neurological examination and a bedside neuropsychological battery known as the Short Cognitive Exam, SCE ^8^, Mini-Mental State Examination (MMSE), the MRC Scale ^6^, and several other pre-existing or adapted clinical scales not directly relevant to current work. Once enrolled, assessments are repeated at domiciliary visits at 4-8 week intervals, depending on rate of clinical change; where patients were progressing more rapidly, visits were at shorter intervals to address clinical need and capture change in condition. All patients were also assessed remotely using the MRC Scale at 2-week intervals.

For the Prion Disease Cognitive Scale development, the study population included all patients with a clinical diagnosis of probable sCJD ^9^ recruited to the NPMC between October 2008 and December 2016 (total n = 431, of which the diagnosis was pathologically confirmed in 231; 54%). For the Prion Disease Motor Scale, this included all patients with a clinical diagnosis of sCJD recruited to the NPMC between July 2013 and December 2016 (n = 168, 78 pathologically-confirmed; 46%). The later inclusion date reflects the time at which additional assessments of cerebellar function were added to the NPMC: the Scale for the Assessment and Rating of Ataxia, SARA ^10^; and the Composite Cerebellar Functional Severity score, CCFS ^11^, components of which became incorporated in the new Motor Scale developed here.

Cognitive and Motor Scale ratio analyses were undertaken using additional assessments from patients with other prion disease aetiologies (inherited and acquired), recruited to the NPMC between October 2008 and December 2016.

### Item bank development

Two “item banks” were created from routinely completed elements of the standardised examination, or other pre-existing scales that could reflect motor or cognitive function. Each examination component, or “item” (e.g. upper limb ataxia, or forward digit span), was documented as a numeric, ordinal score. Data from all individual domiciliary visits were pooled, followed by exploratory cross-sectional analyses of data completeness and patient demographics.

The motor item bank was designed to include items dependent on the integrity of the pyramidal and extrapyramidal motor systems and voluntary motor coordination, which are frequently affected in sCJD ^9^. Additional composite or functional items that could also capture impairment of one or more motor pathways were included, such as eye movements, gait and mobility.

The cognitive item bank was initially composed of all individual items from the Short Cognitive Exam. In addition, to increase face validity, we gave particular weight to items that assessed domains prominently affected in prion disease, which comprises a global dementia with predominant fronto-parietal impairment ^8^. Major challenges of monitoring cognitive function in rapidly-progressive dementias such as sCJD include the attrition rate of patients who are only able to complete one or two longitudinal assessments prior to entering an advanced disease stage ^12^, and patient fatigue, necessitating a brief assessment tool able to measure differences between severely impaired and dysphasic patients. We sought to mitigate this by limiting the final scale to 6 items, and including in the final bank those items with the greatest pooled standardised effect size between two sequential patient assessments, to maximise the ability to stratify patients over the minimum number of assessments.

### Scale development using Rasch analysis

Many of the elements included in each item bank comprised several response categories (“thresholds”). We used the partial credit form of polytomous Rasch analysis (RUMM2030, standard edition) to iteratively refine each item bank into a unidimensional, interval scale reflecting different degrees of composite motor or cognitive dysfunction in sCJD.

Fit to the Rasch model is assessed using a number of approaches ^13^, including: threshold ordering (ensuring that the difficulty of each item’s ordinal scores do indeed progress in the expected sequence); item-trait and item-person interactions (ensuring latent trait invariance and that items themselves are ordered in a hierarchical manner); local dependencies (ensuring that the score in one item does not depend on the score in another, falsely inflating estimated model fit); differential item functioning (ensuring that scale components perform equally regardless of potential confounders such as age, gender or codon 129). We assessed inter-rater reliability by calculating intra-class correlation coefficients (Van de Winckel et al., 2006) for prospective Motor and Cognitive Scale scores for 30 consecutive patients, from independent assessments at the same visit, completed by either a doctor or specialist nurse. Multiple raters were used across this cohort, with different levels of clinical experience.

### Data imputation and further statistical analysis

Rasch-derived scales encapsulate information on both the subject’s ability and the difficulty of all item thresholds, so the “location” of a subject (even if derived from a subset of the scale) defines the probability of their reaching other thresholds ^14^.

We therefore used the Rasch-derived Motor and Cognitive Scales to impute total scale scores where some of the scale items were missing. For the Motor Scale, this comprised all symptomatic patients recruited to the NPMC between October 2008 – June 2013, prior to the introduction of the SARA to the standardised assessment protocol.

For a large proportion of NPMC patients, either the MMSE or SCE was completed, or the former and a subset of the latter (for example, if assessment time was limited, or the patient tired). We used the Cognitive Scale to impute missing data where no more than 2/6 scale items or 10/20 of total possible score were missing. Patients with an MRC Scale score below 5/20 are generally bedbound, have little awareness of surroundings, are unable to use tools and communicate in single words at best ^6^. We therefore imputed a missing Cognitive Scale score of 0 for patients with an MRC Scale score below 5 and MMSE score of 0. To validate this approach, we collected prospective Cognitive and Motor Scale scores in newly-enrolled patients with prion disease. In 30 consecutive sCJD patients with MRC Scale scores below 5/20 (mean 3.0/20, SD ± 0.84), the mean Cognitive Scale score was 0.3/20 (SD ± 0.91).

## Results

### Patient selection for scale development

#### Motor Scale

One hundred and six (106) patients recruited between July 2013 and December 2016 were included in Rasch analysis to develop the Motor Scale (Table 1), comprising 144 individual assessments. Sixty-two (62) additional patients were excluded from analysis due to missing examination data, and review of these patients revealed them to be at advanced neurodisability (maximum group MRC Scale score 4/20), precluding a formal neurological examination beyond assessment of consciousness or limb tone. We reasoned that outcome measures aimed at stratifying motor function in patients at such advanced disability would not be clinically meaningful, nor possible given a dearth of clinical data. All patients were diagnosed as having probable sCJD by consensus criteria ^9^. Forty-eight (48, 45%) underwent post-mortem, which confirmed the diagnosis in all cases.

**Table 1:**
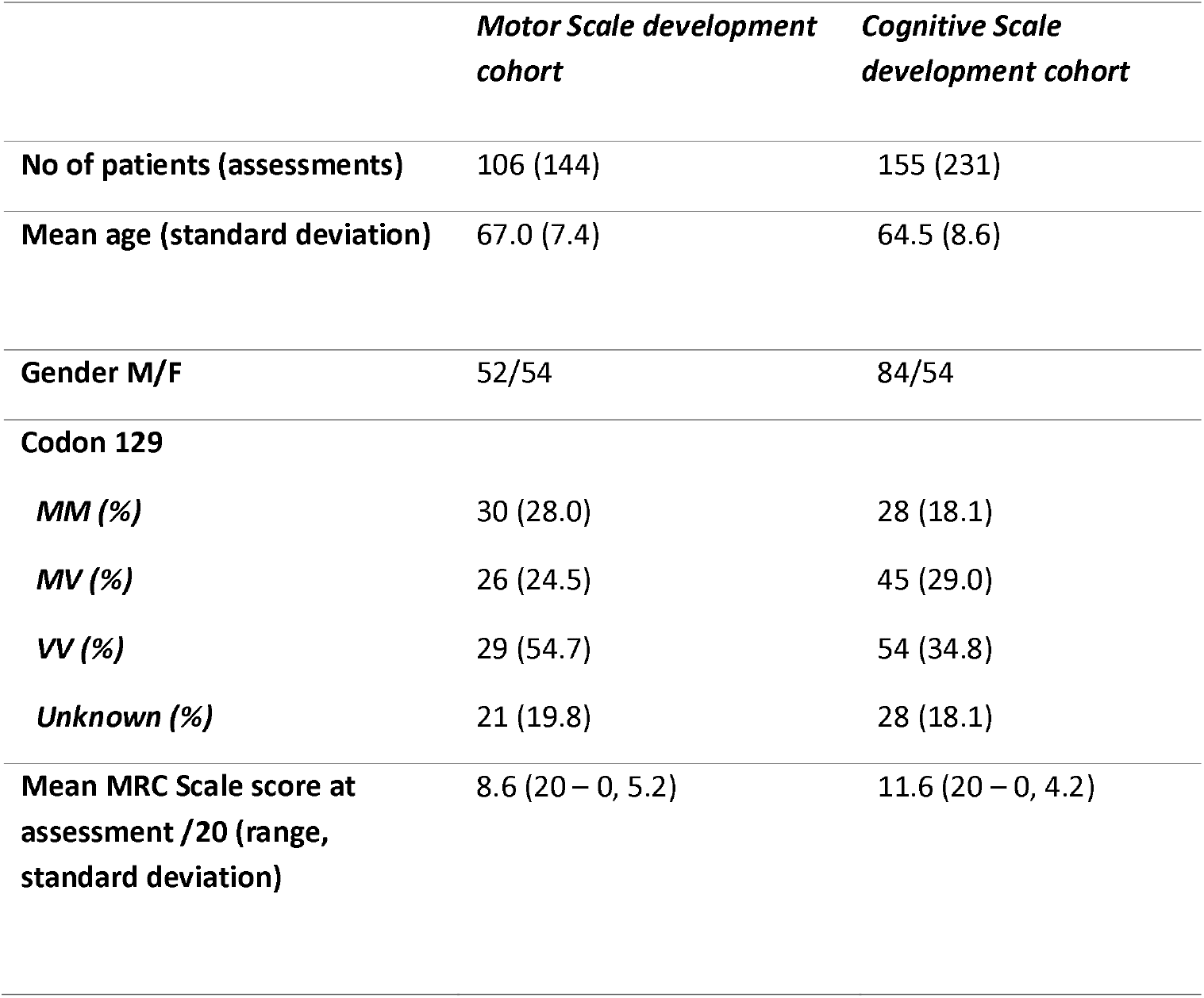
Demographic characteristics for patients included in Motor and Cognitive Scale development cohorts

#### Cognitive Scale

Two hundred and thirty-one (231) assessments from 155 unique patients were included in the Cognitive Scale development (Table 1). Seventy-seven (77, 50%) patients underwent post-mortem examination, which confirmed the diagnosis of sporadic CJD in all cases. Two hundred and seventy-five (275) additional patients were excluded from the scale development cohort, the majority due to advanced disability, which precluded formal cognitive assessment using the SCE.

### Motor Scale development

An initial bank of 20 items was pooled as a single scale, but demonstrated poor fit to the Rasch model. Items were iteratively altered or removed according to threshold order, item and person trait interactions, co-dependency and differential item functioning.

A final, 8-item, 20-point MRC Prion Disease Motor Scale (Motor Scale) (Figure 1) demonstrated good overall fit to the Rasch model. The person separation index was 0.72 (where values above 0.70 are acceptable ^15^) and χ^2^ = 27.2 (df = 24, p = 0.29), without local dependence between items or differential item functioning. A mean item fit residual of -0.28 (SD ± 1.00) and person fit residual of 0.32 (SD ± 0.86) implied both scale items and patients fit the Rasch model. The final Motor Scale included items reliant on pyramidal, extrapyramidal and cerebellar systems, in addition to gait, which is reliant on multiple motor systems. Items that were excluded due to poor ability to discriminate patients at different stages of motor progression included the presence and severity of myoclonus, assessment of limb tone and deep tendon reflexes – concordant with our experience that these aspects of the disease do not necessarily worsen as the disease progresses, and can respond to symptomatic treatments. The intra-class correlation coefficient (ICC, one-way random effects model) was 0.98 (95% CI 0.96 – 0.99), indicating excellent inter-rater reliability.

**Figure 1:**
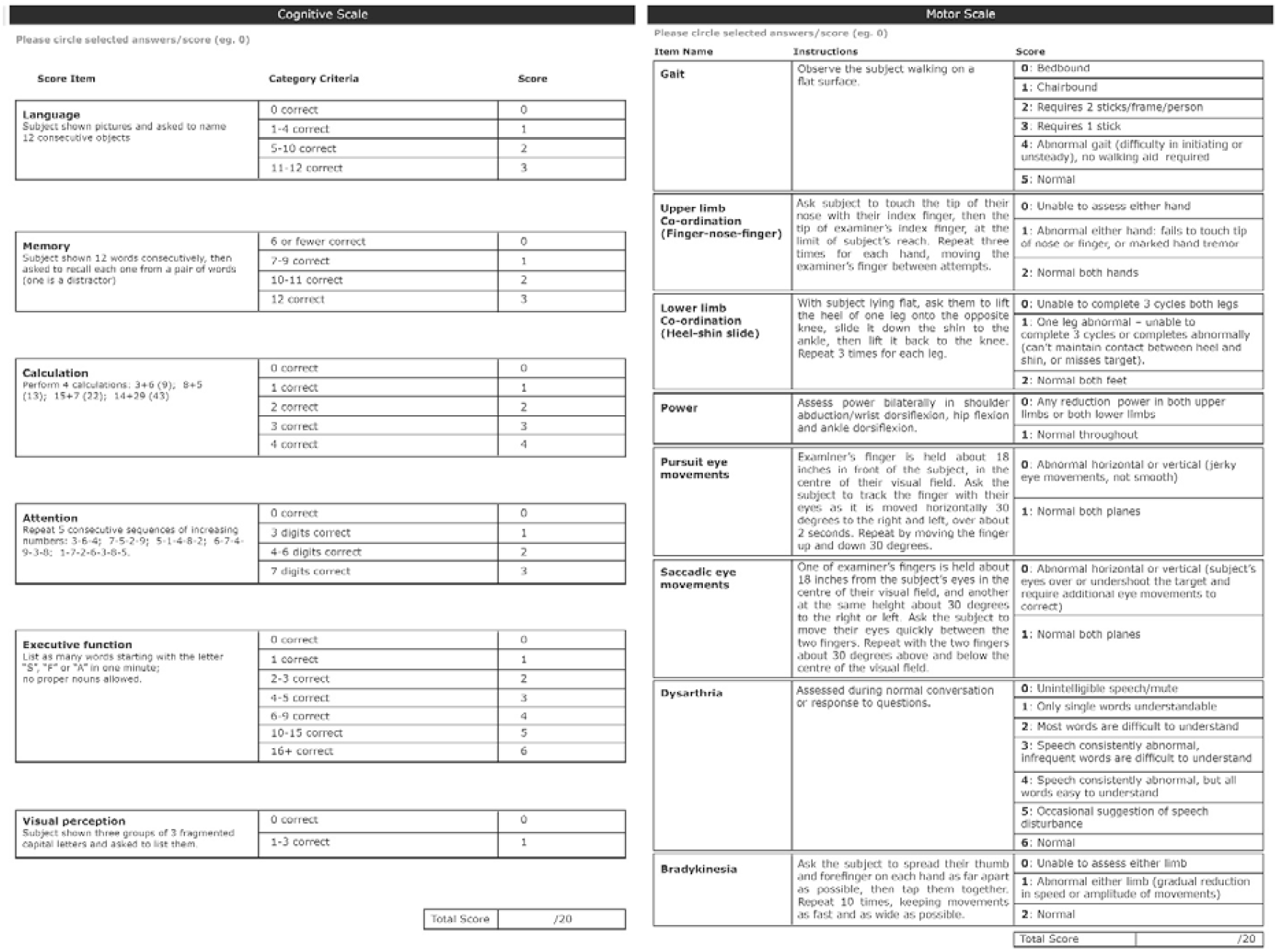
Final Rasch-derived Cognitive and Motor Scales, including instructions and scoring

The Rasch model permits ordering of items and their component thresholds according to their hierarchical difficulty, reflecting composite ability in a desired trait. The final Motor Scale can therefore be used to infer the relative pattern of progression in motor dysfunction for patients with sCJD, as demonstrated in Figure 2. Amongst the earliest motor features in the natural history of sCJD are impaired saccadic and smooth pursuit eye movements, followed by abnormal gait. Conversely, grossly reduced limb power is a relatively late feature.

**Figure 2:**
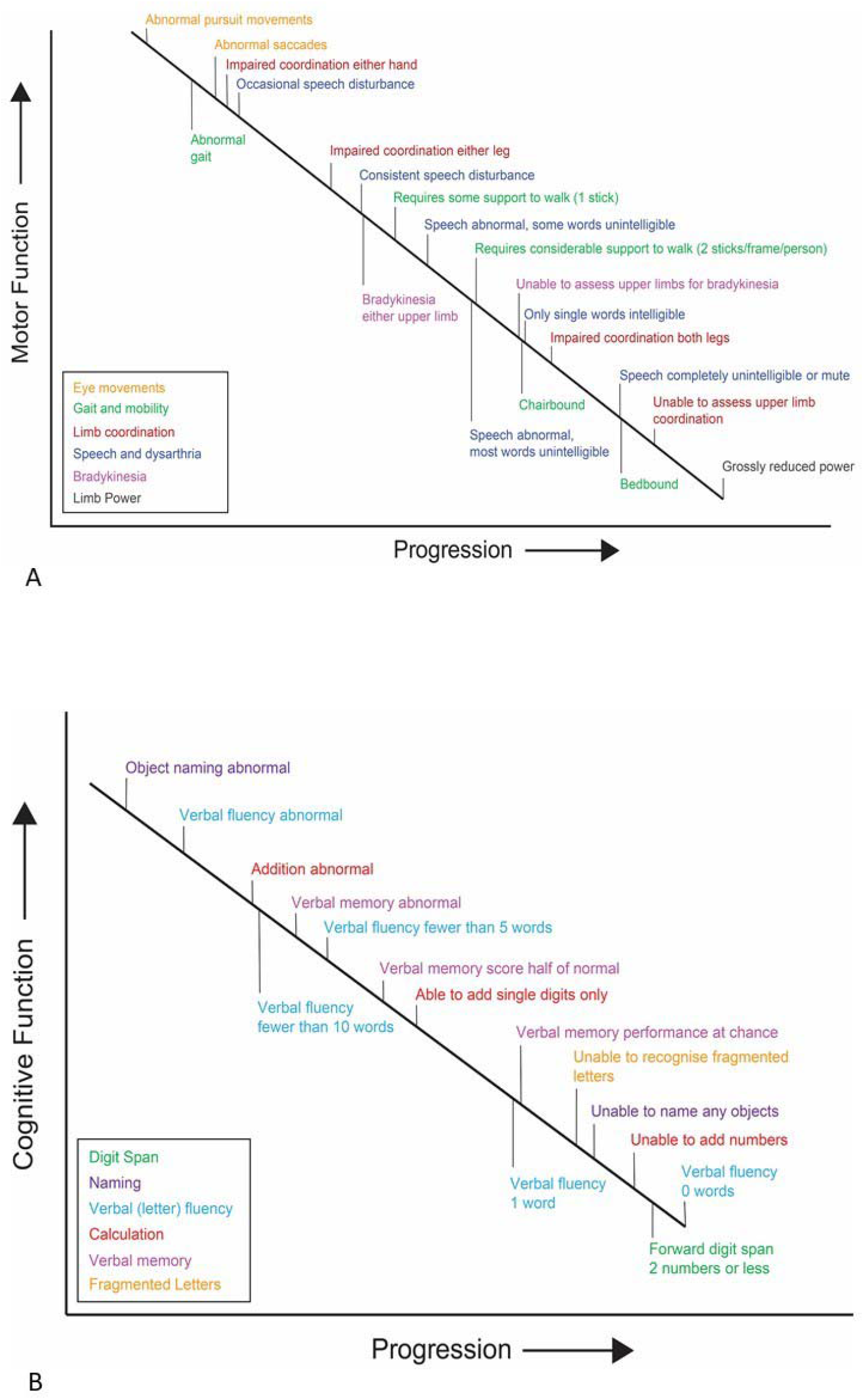
Typical pattern of progressive (A) motor and (B) cognitive dysfunction in prion disease, according to the Rasch model

### Cognitive Scale development

The initial cognitive item bank included a brief set of standardised assessments designed for use in domiciliary visits for patients even with moderately advanced prion disease ^8^. Items included assessment of executive function, language, parietal lobe function, visual perception, recognition memory attention and praxis.

Standardised effect sizes for each item were calculated for 56 patients with two serial assessments, and ranged from 0.55 (calculation) to 0.24 (reading). Spelling and reading items were excluded from the item bank on the basis of low standardised effect sizes, and that the cognitive domains they assessed (parietal lobe function and language, respectively) were also represented by other items. The remaining items were scaled where necessary into ordinal bins to ensure similar total scores for each item. An initial composite scale demonstrated reasonable fit to the Rasch model as judged by mean item and person fit residuals, but inadequate ability to discriminate patients (person separation index 0.66) and no item invariance (χ^2^ = 32.5, df = 18, p = 0.02). The item bank was iteratively refined; all three praxis tasks were removed, having demonstrated poor individual ability to discriminate patients and overall model fit improved. Visual recognition was removed due to local dependence with verbal recognition.

The final MRC Prion Disease Cognitive Scale (Cognitive Scale) comprised 6 items, including measures of executive function, language, parietal lobe function, visual perception, recognition memory and attention/working memory (Figure 1). It displayed good fit to the Rasch model: an acceptable ability to discriminate patients (person separation index 0.74); item invariance (total item χ^2^ = 21.1, df = 18, p = 0.27); good item fit (mean fit residual 0.07, SD ± 0.92) and person fit (mean fit residual -0.45, SD ± 0.99). There were no local dependencies or differential item functioning. The intra-class correlation coefficient was 0.98 (95% CI 0.95 – 0.99).

The Rasch-derived schematic pattern of progressive cognitive dysfunction in sCJD implies earlier deficits in language (naming) and executive function (verbal fluency), with deficits in recognition memory and visual perception occurring later (Figure 2).

### Using Cognitive and Motor Scales to quantitively demonstrate phenotypic heterogeneity

A complete Cognitive and Motor Scale dataset was constructed using three approaches: prospectively-assessed scores from October 2018 to August 2019 (Motor n = 182, Cognitive n = 177); scores calculated in retrospect from items included in the final Rasch-derived scales (Motor n = 293, Cognitive n = 361); imputing missing components of Cognitive and Motor Scale scores as outlined in Methods (Motor n = 1403, Cognitive n = 833). In total, 528 patients with sCJD had at least one Cognitive or Motor Scale score.

Individual Cognitive and Motor Scale longitudinal patient trajectories suggested an approximately linear decline in function over time in most patients with sporadic CJD, and a decrease in scale score between enrolment and a subsequent assessment (Figure 3). Higher Cognitive or Motor Scale scores at enrolment were associated with longer subsequent survival in patients with sCJD (Spearman’s rho: Cognitive Scale = 0.46, p < 0.00001; Motor Scale = 0.51, p < 0.00001).

**Figure 3:**
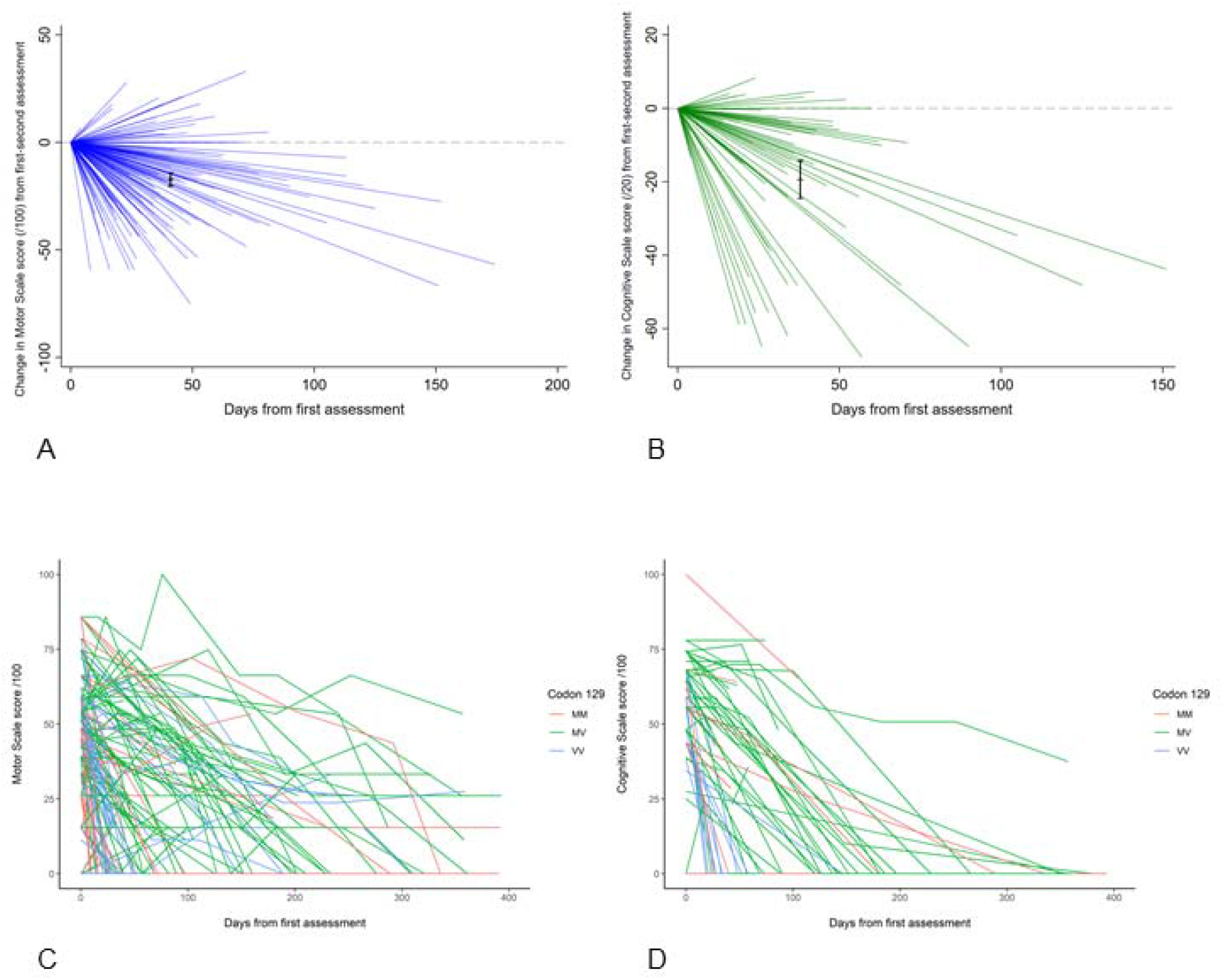
Change in Motor Scale (A) and Cognitive Scale (B) score between first (anchored to score of 0) and second assessment for individual patients with sCJD, /100; reference line indicates no change between assessments, mean change (95% CI) and mean follow-up date in black. Spaghetti plots of individual sCJD patient trajectories for Motor (C) and Cognitive (D) Scale scores over multiple assessments, grouped by PRNP Codon 129 polymorphism.

We estimated a simple least-squares linear regression model of each patient’s first Cognitive and Motor Scale score with paired MRC Scale scores to evaluate the proportion of variance in Cognitive and Motor function explained by overall disease progression. Changes in the MRC Scale explained 72% of variance in Cognitive score (model *F* (1, 402) = 1052.4, p < 0.00001) and 80% of variance in Motor score (model *F*(1, 496) = 1961.0, p < 0.00001). We estimated that a Cognitive Scale score of 1/20 was reached at MRC Scale score = 3.9 (95% CI 3.6 – 4.3) and Motor Scale score 1/20 at MRC Scale score 3.5 (95% CI 3.2 – 3.7), indicating expected floor effects at this level, at which point patients are bedbound, unable to verbalise and without awareness of their surroundings ^6^.

Finally, we explored the potential for paired Cognitive and Motor Scale scores to quantitatively reflect phenotypes between prion disease subtypes, which result in different patterns of cognitive and motor dysfunction ^4^. We calculated Cognitive/Motor Scale score ratios (CM ratio) for patients with at least one pair of assessments (total n = 570, sCJD n = 435; Table 2), where a CM ratio of greater than 1 indicates greater relative motor dysfunction; and less than 1, greater relative cognitive dysfunction. We compared the first available CM ratio in patients with an MRC Scale score greater than 4/20, for patients according to prion disease subtype (Figure 4), and across codon 129 genotype in patients with sCJD (Figure 5).

**Table 2:** Cognitive / Motor ratio cohort demographics

|  | All Patients |  |  |  |  | sCJD Patients |  |  |  |  |
| --- | --- | --- | --- | --- | --- | --- | --- | --- | --- | --- |
| Variable | No of patients<br>(Total Paired Records) | No with MRC (Total Paired Records) | Mean 1<sup>st</sup> Assessment MRC ± SD (95% CI) | Mean 1<sup>st</sup> Assessment Motor100* ± SD (95% CI) | Mean 1<sup>st</sup> Assessment Cog100* ± SD (95% CI) | No of patients (Records) | No with MRC (Records) | Mean 1<sup>st</sup> Assessment MRC ± SD (95% CI) | Mean 1<sup>st</sup> Assessment Motor100* ± SD (95% CI) | Mean 1<sup>st</sup> Assessment Cog100* ± SD (95% CI) |
| At least one paired Motor & Cognitive scale score | 570 (1286) | 545 (1169) | 7.4 ± 6.9<br>(6.8 – 8.0) | 30.3 ± 29.4<br>(27.9 – 32.8) | 27.0 ± 31.8<br>(24.4 – 29.7) | 435 (728) | 427 (690) | 6.1 ± 6.1<br>(5.5 – 6.7) | 24.6 ± 26.0<br>(21.1 – 27.1) | 22.0 ± 29.5<br>(19.2 – 24.8) |

\*Motor and Cognitive scale scores transformed to scores /100 according to the Rasch model.

**Figure 4:**
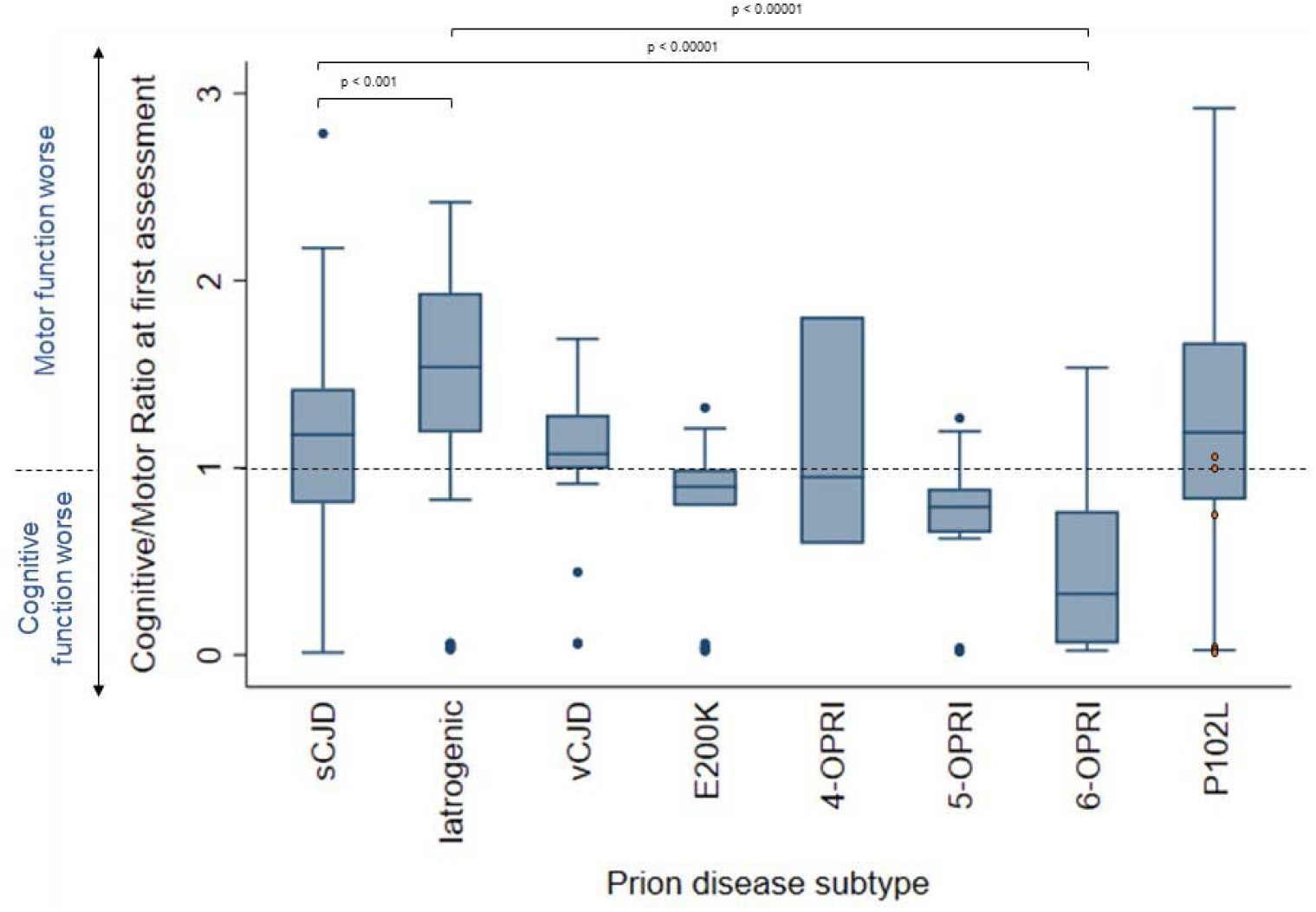
Cognitive / Motor scale score ratios (CM ratios) at first assessment for different prion disease aetiologies. Displayed are median scores, interquartile ranges (IQR, boxes) and limits of Q1/Q3 + 1.5xIQR (whiskers). Subtypes were compared using the Kruskal Wallis test for non-parametric data, followed by ad hoc pairwise comparison with Dunn’s test, adjusted for multiple comparisons. sCJD = sporadic CJD; iatrogenic = iatrogenic CJD; vCJD = variant CJD; inherited prion disease (E200K mutation); inherited prion disease (4-OPRI mutation); inherited prion disease (5-OPRI mutation); inherited prion disease (6-OPRI mutation); inherited prion disease (P102L mutation) – patients with a CJD phenotype ^32^ are marked as orange circles. Differences shown here illustrate the face validity of our scales.

**Figure 5:**
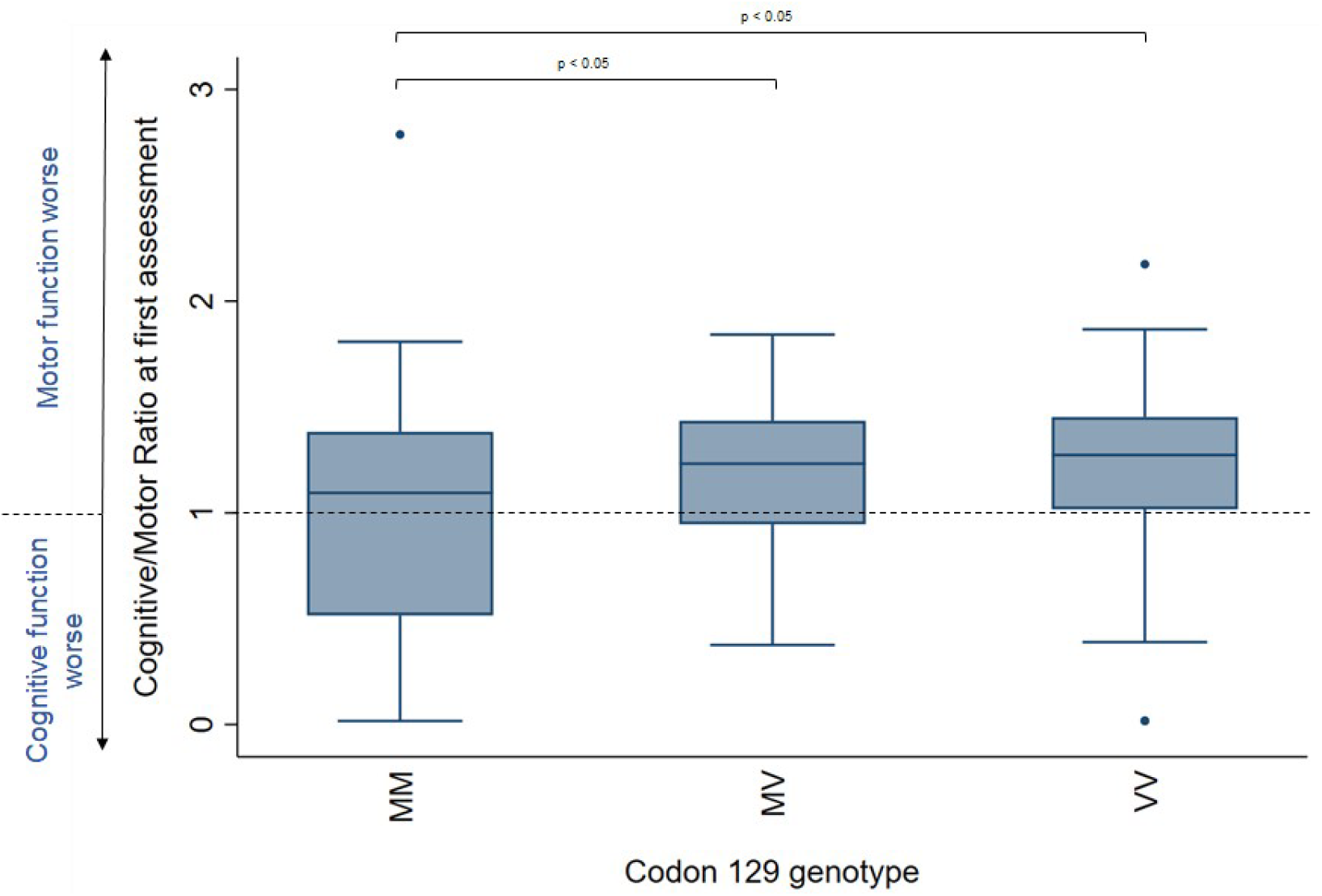
Cognitive / Motor Scale score ratios (CM ratios) at first assessment for sCJD patients according to codon 129 polymorphism. Displayed are median scores, interquartile ranges (IQR, boxes) and limits of Q1/Q3 + 1.5xIQR (whiskers). Subtypes were compared using the Kruskal Wallis test for non-parametric data, followed by ad hoc pairwise comparison with Dunn’s test, adjusted for multiple comparisons.

Patients with iatrogenic CJD (iCJD) due to treatment with contaminated cadaveric growth hormone typically present with early cerebellar ataxia and dysarthria, but relatively preserved cognition ^16^. This is reflected in the median CM ratio of patients with iCJD, which is significantly greater than those with sporadic CJD (Kruskal Wallis/Dunn’s post-hoc pairwise comparison p < 0.00001). Conversely, patients with inherited prion disease caused by the six octapeptide repeat mutation (6-OPRI) manifest a striking cortical dementia, with onset of motor system dysfunction occurring much later ^2, 17^ – as evidenced by a low CM ratio.

Amongst patients with sporadic CJD, the median CM ratio differed according to codon 129 genotype. The median CM ratio for patients homozygous for methionine at codon 129 was significantly lower than those heterozygous or valine homozygous, indicating greater relative motor dysfunction in the latter two groups (Figure 5). Twenty-six (26) patients were classified by post-mortem molecular PrP^Sc^ typing under the London classification ^3^ (Table 3), which can be broadly compared to another commonly used classification system ^4, 18^. The mean of first available CM ratio for patients with the *PRNP* 129 MM genotype and PrP^Sc^ type 2 (London classification; Parchi MM1) was 0.94 (SD ± 0.90), indicating predominant cognitive dysfunction at first assessment, and for *PRNP* 129 VV genotype and PrP^Sc^ type 3 (Parchi VV2), 1.41 (SD ± 0.36), indicating greater relative motor dysfunction.

**Table 3:** PrP^Sc^ type, PRNP genotype and CM ratios for selected sporadic CJD patients

| PrP<sup>Sc</sup> Type (London Classification*) | PRNP genotype | No of patients | Mean CM Ratio (SD) |
| --- | --- | --- | --- |
| 2 | MM | 3 | 0.94 (0.90) |
| 2 | MV | 3 | 0.83 (0.15) |
| 2 | VV | 0 | - |
| 3 | MM | 3 | 0.59** (0.23) |
| 3 | MV | 8 | 1.13 (0.33) |
| 3 | VV | 8 | 1.41** (0.36) |

\* Strain typing undertaken on post-mortem frontal cortex samples, as described in <sup>33</sup>.
\*\* Pairwise comparisons of mean CM ratios were significant between PrP<sup>Sc</sup> type 3 MM and VV genotypes (Kruskal Wallis test for non-parametric data, followed by ad hoc pairwise comparison with Dunn's test, adjusted for multiple comparisons, $p < 0.05$ ).

## Discussion

We have developed two validated, disease-specific, clinician examination-based rating scales for patients with sCJD, reflecting disease progression in cognitive and motor systems. The scales were developed using item-response modelling and have favourable statistical properties. Both scales are short, in that they can be completed in a few minutes, avoid jargon or abstract scoring systems, and require no special equipment other than images on cards for the cognitive tests. They use items commonly assessed as part of a routine neurological and bedside cognitive examination. These scales complement the functionally-orientated MRC Scale and have several potential uses.

The ability to objectively quantify motor and cognitive dysfunction in prion disease should add valuable clinimetric tools to assess patients both in formal clinical trials and routine neurological practice. Differences in the speed and extent of motor versus cognitive dysfunction can help to stratify patient phenotypes that progress at different rates ^19^, and thus provide more accurate prognostic information for patients and their families. Moreover, documenting a patient’s motor or cognitive function using these scales will allow inferences regarding the likely future sequence and pace of clinical change, and thus allow more effective planning of future care needs and information provision ^20^. Additionally, informal pilot use of both scales via telemedicine has proven feasible with the support of a local clinician able to examine the patient, and could allow remote assessment in future practice.

Previous efforts have been made to develop or use system-specific clinical rating scales in the setting of prion disease therapeutic trials. These have focussed predominantly on the ordinal assessment of individual neurological examination features, such as degree of ataxia or presence of frontal release signs, or total accretion of neurological signs ^21, 22^. Similar examination components, or items, comprised the item banks used for development of the Motor and Cognitive Scales. However, these measures suffer with the issues that warranted the use of Rasch methodology to create the Motor and Cognitive Scales: they are not validated as measuring a single construct, nor as a composite or true interval score, and are by definition dependent on the population in which they are measured ^7^. The same problems affect common cognitive assessments of patients with prion disease. These have typically included the Mini-Mental State Examination (MMSE), which does not assess executive function ^23^ – commonly impaired in prion disease ^8, 24^ – and the Clinical Dementia Rating (CDR) scale, which is time-intensive to administer, requires training, and is heavily dependent on carer interview^25^.

The key elements of each final scale were broadly consistent with clinical signs and symptoms most prominent in prion disease. A large portion of the Motor Scale involved direct assessment of cerebellar function, which is impaired at presentation in the majority of patients with sCJD; this contrasts with extrapyramidal dysfunction, which constitutes a small portion of the Motor Scale and is generally observed later in the disease course ^26^. We found that altered limb tone or deep tendon reflexes are not useful measures to stratify motor dysfunction in prion disease. Interestingly, despite its textbook association with prion disease and observation as part of most clinical phenotypes ^26^, myoclonus was also not useful in measuring motor dysfunction. This may relate to its binary classification (present, not present) despite efforts to grade severity, and its spontaneous remission as the disease progresses in some cases, or improvement in some patients receiving the highly effective symptomatic treatments that are available ^27^.

The profile of progressive cognitive impairment in prion disease implied by the Cognitive Scale supports a leading role for executive dysfunction with expressive language impairment, with recognition memory a less prominent component ^8^. The Heidenhain variant phenotype is a well-documented presentation of sporadic CJD involving higher order visual dysfunction at onset; visual perception was affected late in the Cognitive Scale, and was a small contributor to progression. This is likely to reflect the strength and limitation of Rasch-derived scales: by definition, scales are a composite of the most useful items to reflect the entire span of the desired trait – thus, items that assess rare symptoms or signs have little value in stratifying most patients, and are likely to be excluded. Although one of the most commonly discussed clinical phenotypes, the Heidenhain variant affects only about 5% of patients with sporadic CJD ^28^.

There are other limitations to this work. Polymorphism at codon 129 of the prion protein gene is the major known modifier of disease progression in sporadic CJD and many other prion disease subtypes ^29, 30^. As with most clinical studies of prion disease, codon 129 methionine homozygotes are under-represented in this study, as their generally shorter disease course means they are often assessed at advanced stages of neurodisability. Although comprising a smaller proportion of the scale development cohorts than reflected by epidemiological surveillance of sCJD cases, we feel methionine homozygotes constituted a sufficient number of assessments to allow development of balanced scales. Rasch methodology identifies shared variables that reflect an underlying trait of interest (motor or cognitive function) that should perform equally well regardless of the subpopulation (for example, codon 129) assessed. The absence of differential performance of the scales in different codon 129 groups reflects this property, and suggests that a lower proportion of methionine homozygotes has not biased their development.

PrP^Sc^ typing was only available for a small subset of patients with sporadic CJD. Interestingly however, the mean Cognitive/Motor ratio does appear to stratify the predominant phenotypes described in association with certain PrP^Sc^ types/PRNP genotypes (Table 3): for example, type 3 PrP^Sc^ with VV or MV at codon 129 ^3^, or type 2 PrP^Sc^ by the Parchi classification ^4^, present with striking ataxia; compared with type 2 or 3 PrP^Sc^ with MM at codon 129 (MM1/MM2C according to the Parchi classification), in which cognitive dysfunction is the dominant early feature ^4^. It should be noted that PrP^Sc^ typing was available for a limited number of cases, and pairwise comparisons of mean Cognitive/Motor Ratios was significant for only type 3 PrP^Sc^ with MM or VV genotypes (Table 3); further analysis with a larger sample size is required to formally test this hypothesis.

Finally, both Motor and Cognitive Scales demonstrated floor effects, at mean MRC scores of 3.5 and 3.9/20, respectively. Patients at this advanced level of disability are generally bedbound, with limited verbal or physical communication ^6^, and prognosis at this disease stage very limited ^31^. Given the intended scale use in routine neurological practice and future clinical trials, the ability to identify and stratify change in motor or cognitive function is likely to be extremely challenging, and of limited use during this phase.

We used Rasch methodology to develop two novel clinical rating scales assessing progressive motor and cognitive dysfunction in prion disease. These address many of the obstacles involved in assessing clinical change in rapidly-progressive, multisystem dementias, and may prove useful in future clinical trials, prognostic modelling and feedback as part of routine clinical care. A similar approach could be employed for other rapidly-progressive dementias or comparable neurological conditions.

## Data Availability

Anonymised data available on request to the corresponding author.

## Acknowledgements

We thank all the individuals, their caregivers and families who took part in the National Prion Monitoring Cohort, and the UK clinicians and National CJD Research and Surveillance Unit for referring patients. We thank all current and past colleagues at the National Prion Clinic who undertook the clinical assessments utilised as part of this research. We thank Richard Newton, who kindly assisted in preparing figures, and Joanna Field and Sarah Mazdon for support with data collation.

The National Prion Monitoring Cohort study was funded by the Department of Health and Social Care (England), Medical Research Council (core funding to MRC Prion Unit), and University College London Hospitals/University College London National Institutes for Health Research Biomedical Research Centre. Akin Nihat is supported by a Medical Research Council Clinical Research Training Fellowship (grant number MR/P019862/1). Tze How Mok is supported by a Fellowship award from Alzheimer’s Society, UK (grant number 341 (AS-CTF-16b-007)).

## Author Contributions

Conception and design of study: A.N., S.M., P.R., J.C., T.H.M., A.T., D.C. All authors contributed to data acquisition and analysis: A.N., S.M., P.R., T.H.M., A.T., H.O., K.M., V.O., S.T. Drafting of the manuscript: A.N., S.M. All authors have read and approved the final manuscript.

